# Artificial-Intelligence-Enabled Early Malnutrition Risk Assessment Tools for Elderly Trauma Patients in Intensive Care Units

**DOI:** 10.64898/2026.04.26.26351765

**Authors:** Xinhang Wei, Xiang Cao, Jianguo Hou, Qi Wang

**Author notes:** Corresponding author: Qi Wang.

## Abstract

**Background & Aims:** Accurate assessment of clinical malnutrition using anthropometric and functional indicators could improve the care of elderly trauma patients in intensive care units (ICUs). This study aimed to develop an AI-driven malnutrition assessment toolbox based on a minimal set of clinically feasible indicators.

**Methods:** Multiple machine learning models, including logistic regression, support vector machines, k-nearest neighbors, decision trees, random forests, XGBoost, and neural-network-based ensemble models, were developed using different indicator configurations from a clinically collected patient dataset. Models were trained using baseline and longitudinal measurements to predict malnutrition risk. SHAP analysis was used to interpret the importance of selected indicators.

**Results:** Baseline (Day 1) data alone did not provide a reliable prediction, whereas longitudinal measurements substantially improved performance. Models based on a minimal indicator set, including bilateral mid-upper arm circumference, calf circumference, and key static variables, outperformed models using the full indicator set. Tree-based methods consistently outperformed linear and distance-based models, with the three-time-point XGBoost achieving the best individual performance. Neural-network-based ensemble models further improved predictive stability. The best overall performance was achieved by the ensemble model using the minimal indicator set from Day 1 and Day 3. SHAP analysis confirmed the importance of the selected indicators.

**Conclusions:** This AI-driven toolbox provides an efficient and clinically feasible approach for early malnutrition assessment in elderly trauma patients in the ICU. Its strong performance with a minimal indicator set supports its potential for integration into clinical workflows and future digital twin systems for intelligent nutritional management.

## 1. Introduction

As the population rapidly ages in many parts of the world, trauma has become a major cause of death among the elderly [1,2]. For example, China’s aging population has resulted in a corresponding increase in the proportion of elderly trauma patients [3]. These patients often face various nutritional challenges, including inadequate energy intake, diabetes, obesity, and sarcopenia, which lead to a much higher rate of malnutrition among elderly trauma patients compared to younger individuals [4].

Studies have demonstrated a strong correlation between malnutrition and adverse clinical outcomes in numerous clinical conditions [5-8]. Some studies also indicate that elevated body fat percentage and reduced skeletal muscle mass index are closely tied to the nutritional risk in elderly trauma patients as well [9].

Malnutrition constitutes a significant challenge in the recovery, prognosis, and treatment of elderly individuals who have sustained traumatic injuries. It not only weakens the immune system and delays wound healing but may also increase the risk of infection and other complications, interrupting standard treatment processes. In most cases, it can prolong hospitalization and increase medical expenses [10-16]. In severe instances, it may lead to death [17]. Therefore, quickly identifying and accurately assessing malnutrition in elderly trauma patients is essential to improve the treatment of underlying diseases and clinical outcomes.

Advances in artificial intelligence (AI) in healthcare and medicine have brought transformative changes to the field [18-20]. However, there has been no systematic effort to develop AI-enabled tools to quickly and accurately evaluate malnutrition in elderly trauma patients using clinically available data. This study aims to create a set of AI-enabled, data-driven diagnostic tools to assess malnutrition in elderly trauma patients, built on a small, clinically available dataset. These tools take the input from a minimal set of anthropometric and functional indicators that adhere to the Global Leadership Initiative on Malnutrition (GLIM) criteria. The outcome variable is derived directly from GLIM-based clinical assessment, with patients classified as either non-malnourished (Class 0) or malnourished (Class 1), providing a clinically grounded binary label for model training and evaluation.

While GLIM provides a clinically validated framework for malnutrition diagnosis, its practical implementation faces several inherent challenges. Diagnosis under GLIM requires the concurrent satisfaction of at least one phenotypic criterion (e.g., unintentional weight loss, low BMI, or reduced muscle mass) and at least one etiologic criterion (e.g., reduced food intake, malabsorption, or disease-related inflammation). Although comprehensive, this multi-dimensional framework depends heavily on the availability of longitudinal and heterogeneous clinical data, many of which are frequently incomplete or difficult to obtain in routine practice. Prior weight history is often unavailable or unreliable, dietary intake assessments are inherently subjective, and inflammatory or imaging-based indicators may not be consistently measured, particularly in resource-limited or emergency settings. Beyond data availability, the assessment process relies substantially on clinician judgment, introducing inter-rater variability and limiting scalability, standardization, and automation across clinical environments.

These limitations highlight a critical gap between the diagnostic rigor of GLIM and its operational feasibility at the bedside. This study addresses this gap by developing a machine learning–based surrogate framework that predicts GLIM-defined malnutrition risk using a minimal set of routinely collected, readily measurable clinical indicators—specifically, bilateral mid-upper arm circumference (MAC), calf circumference (CC), and key static variables. Rather than replacing or redefining the GLIM standard, the proposed approach operationalizes it in a practical, data-driven form. Crucially, the framework enables early risk stratification before full GLIM criteria can be formally evaluated, supporting timely clinical intervention during the critical early stages of hospitalization. By delivering automated, reproducible, and scalable predictions from compact anthropometric inputs, this toolbox transforms a retrospective diagnostic standard into a forward-looking, clinician-friendly risk assessment tool with strong potential for seamless integration into routine clinical workflows and future intelligent healthcare systems [29,30].

This toolbox comprises several foundational machine learning models, including logistic regression, SVM, KNN, decision trees, random forests, XGBoost, and neural-network-based ensemble models, trained on clinically collected patient data from elderly trauma patients. It also includes a selected foundational data processing and engineering module to produce AI-ready datasets for training the machine learning models. At clinical and hospital levels, the models can be readily retrained on new or updated datasets from trauma patients through transfer learning. The toolbox provides a ranking of model performance and options for clinical users to choose their preferred model. This toolbox can then be used either as a clinical aid to clinicians or further developed into a digital twin component for trauma patients in the future.

In this paper, we begin with a dataset from a small cohort of elderly patients, consisting of anthropometric and functional indicators and malnutritional assessment results adherent to the Global Leadership Initiative on Malnutrition (GLIM) criteria. We show that baseline (Day 1) measurements of the indicators alone are insufficient for reliable prediction, whereas incorporating longitudinal data markedly improves performance. We then search for a minimal set of indicators with the greatest discriminant power that is also easy to measure under an emergency. We find that the set of bilateral mid-upper arm circumference (MAC), calf circumference (CC), and 6 key static variables (defined in the next section) provides superior assessment outcomes. The assessment tools built with these indicators outperform those built with the full set of indicators. We then compare the performance of all the machine learning models and find that tree-based models consistently outperform linear and distance-based approaches, with the three-time-point XGBoost achieving the strongest overall performance. To further improve the performance of individual machine learning models, we develop a neural-network-based ensemble approach that produces models from the ensemble of the best performers. As a result, we show that neural network–based ensemble models further improve predictive stability within the same indicator configuration, and the ensemble model based on the minimal indicator set from Day 1 and Day 3 captures most of the predictive signal and provides the best overall assessment of malnutrition, thereby offering an efficient and clinically feasible solution.

## 2. Data acquisition and processing

This study is a multi-center prospective cohort study aimed at enrolling trauma patients aged ≥60 years old. The study excluded patients transferred from other hospitals, those who suffered from non-mechanical trauma (such as burns, frostbite, electric injuries, acid-base injuries, poisonings, animal bites, and insect stings), and patients hospitalized for fewer than 3 days. The GLIM tool was employed to assess patients’ nutritional status using a two-step approach: first, a nutrition risk screening was conducted using the Nutrition Risk Screening 2002 (NRS-2002) and Mini Nutritional Assessment Short Form (MNA-SF); subsequently, phenotypic and etiological indicators were collected on the 1st, 3rd, and 7th days of hospitalization. Among the phenotypic indicators, mid-upper arm circumference (MAC), calf circumference (CC), triceps skin-fold thickness (TSF), and handgrip strength (HGS) were identified as key metrics for evaluating muscle mass. Data imputation and augmentation models were employed to fill in missing entries and generate synthetic data for machine learning, given that the ground-truth dataset comprises only 269 patients.

This study was approved by the Ethics Committee of Sichuan Academy of Medical Sciences & Sichuan Provincial People’s Hospital on June 19, 2023 (Ethical Approval No.: Yan-2023-277) and has been registered with the Chinese Clinical Trial Registry (Registration No.: ChiCTR2300074858). All enrolled patients provided written informed consent prior to participation. The research was conducted in strict accordance with Good Clinical Practice guidelines, relevant national laws and regulations, and the Declaration of Helsinki.

### 2.1 Inclusion and exclusion criteria

We list the inclusion and exclusion criteria below.

<u>Inclusion criteria</u>:

a. Age ≥ 60 years old.
b. Patients initially admitted to the hospital for trauma.
c. Patients are expected to stay in the hospital for up to 3 days or longer.

<u>Exclusion criteria</u>:

a. Patients suffer from burns, frostbite, and electric shock injuries, those with acid and alkali burns and poisoning, and individuals bitten by animals or insects.
b. Patients who have been hospitalized at other facilities.
c. Patients either participated in other studies or declined to participate in this study.

### 2.2 Data analysis

The initial dataset consisted of 269 patients. Eight anthropometric and functional indicators were used to characterize patients’ nutritional status, including Left mid-upper arm circumference (MAC_L), Right mid-upper arm circumference (MAC_R), Left calf circumference (CC_L), Right calf circumference (CC_R), Left triceps skinfold thickness (TSF_L), Right triceps skinfold thickness (TSF_R), Left hand grip strength (HGS_L), and Right hand grip strength (HGS_R). These dynamic measurements were collected at three consecutive time points (Day 1, Day 3, and Day 7). In addition to longitudinal anthropometric and functional indicators, several static baseline indicators were recorded to capture demographic and injury-related characteristics, including age group, sex, injury location, injury type, and status of chronic and acute diseases. These static indicators were incorporated to provide complementary contextual information for subsequent modeling and risk stratification.

Table 1 presents the missing data rates for each dynamic indicator across the three points. Overall, most anthropometric measurements exhibited relatively low levels of missingness on Day 1 and Day 3, typically below 10%. Handgrip strength measurements showed slightly higher rates of missingness at these early time points, reaching approximately 12–13%. A substantial increase in missingness was observed on Day 7, with most indicators showing missing rates of 37% to 42%.

**Table 1.** Data-missing rate on days 1, 3, and 7 across all indicators.

| Indicator | Day 1 | Day 3 | Day 7 | Overall |
| --- | --- | --- | --- | --- |
| MAC_L | 6.32% | 6.69% | 37.55% | 16.85% |
| MAC_R | 5.20% | 5.95% | 37.92% | 16.36% |
| CC_L | 5.58% | 6.32% | 38.66% | 16.85% |
| CC_R | 2.60% | 3.72% | 37.17% | 14.50% |
| TSF_L | 7.43% | 7.81% | 38.66% | 17.97% |
| TSF_R | 5.95% | 6.69% | 39.03% | 17.22% |
| HGS_L | 11.90% | 13.01% | 41.64% | 22.18% |
| HGS_R | 11.90% | 12.64% | 42.01% | 22.18% |

When considering all time points, the overall missing-data rate across the dynamic indicators was approximately 17.29%. Circumference-based and skinfold-based measurements (MAC, CC, and TSF) exhibited moderate overall missing rates, generally between 14% and 19%, while handgrip strength measurements had slightly higher overall missing rates of approximately 21–22%. This correlates with the difficulty of obtaining measurements from trauma patients in emergency situations.

In addition to the dynamic measurements, 6 static categorical indicators were collected to describe baseline patient characteristics, as shown in Table 2. These indicators included injury location (7 categories), injury type (4 categories), chronic and acute disease status, sex, and age group (3 categories). Injury location and injury type displayed uneven categorical distributions, with certain categories containing substantially fewer samples. Chronic disease was present in 227 patients, while acute disease was relatively rare (18 patients). Sex and age group were more evenly distributed, although the third age group contained fewer cases. These static indicators were incorporated to provide complementary baseline information for subsequent modeling analyses.

**Table 2.** Distribution of Static Categorical Indicators.

| Indicator | Category | Count |
| --- | --- | --- |
| Injury location | 1 | 27 |
|  | 2 | 138 |
|  | 3 | 10 |
|  | 4 | 18 |
|  | 5 | 5 |
|  | 6 | 34 |
|  | 7 | 37 |
| Injury type | 1 | 209 |
|  | 2 | 19 |
|  | 3 | 23 |
|  | 4 | 18 |
| Chronic disease | Yes (1) | 227 |
|  | No (0) | 42 |
| Acute disease | Yes (1) | 18 |
|  | No (0) | 251 |
| Sex | 0 | 176 |
|  | 1 | 93 |
| Age group | 1 | 136 |
|  | 2 | 114 |
|  | 3 | 19 |

Figure 1 presents the correlation matrices of the eight anthropometric and functional indicators on Day 1 and Day 3, respectively. Overall, the correlation structures between the two points are highly consistent, with only minor variations in magnitude. Most body-size–related indicators (MAC, CC, and TSF) demonstrate moderate-to-strong positive correlations with each other, generally ranging from 0.60 to 0.70 across time points. As expected, bilateral measurements exhibit very strong correlations. Left and right mid-upper arm circumference show correlations above 0.97, while left and right calf circumference demonstrate correlations around 0.97–0.98. Similarly, triceps skinfold thickness measurements on both sides are highly correlated (r ≈ 0.98) and left– and right-hand grip strength exhibit strong bilateral agreement (r ≈ 0.92–0.93). These findings confirm strong internal consistency within paired anatomical measurements. In contrast, handgrip strength shows relatively weak correlations with anthropometric indicators. Correlation coefficients between grip strength and circumference or skinfold measures generally fall within the 0.20–0.35 range, indicating that functional capacity captures complementary information beyond structural body-size indicators. Comparing Day 1 and Day 3, the overall correlation patterns remain stable, with slight increases in certain associations over time. This temporal consistency suggests that the relationships among nutritional indicators are structurally stable across the early stages of hospitalization. However, the observed variability in correlation strength suggests that temporal dynamics should still be accounted for when modeling longitudinal cohort data.

**Figure 1.**
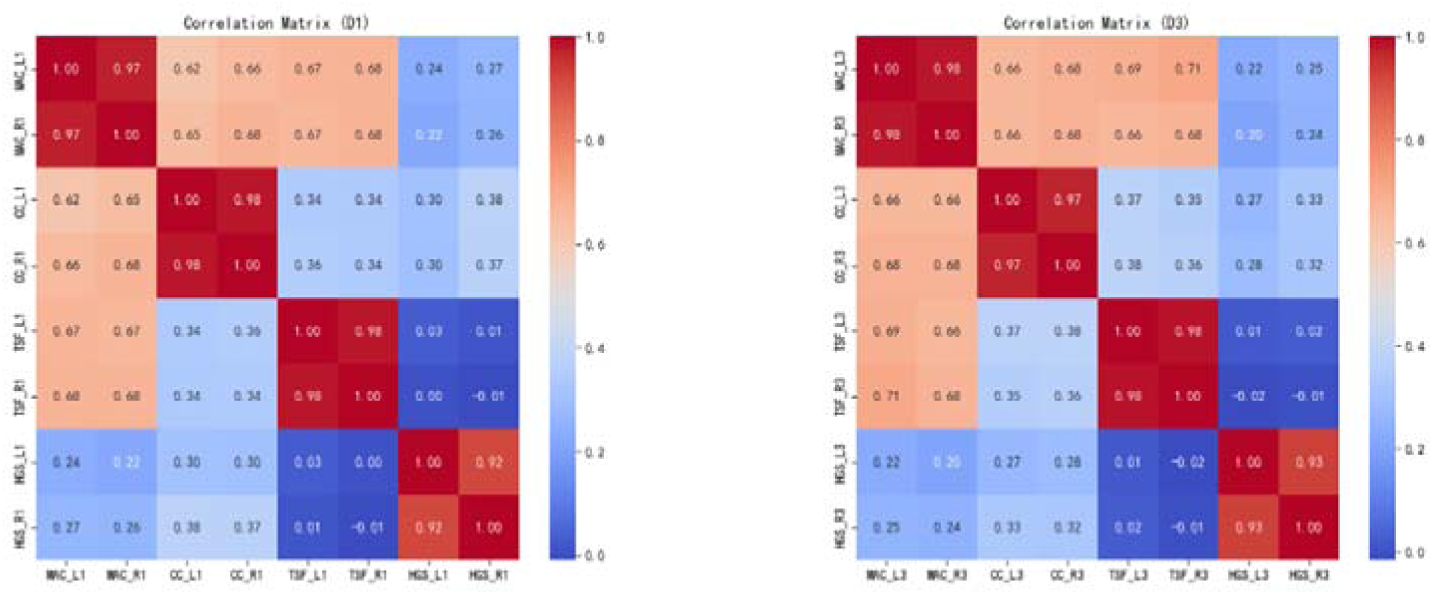
Correlation analysis among all indicators for malnourished (1) and non-malnourished (0) groups on Day 1 and Day 3, respectively. There is no statistically significant difference in the correlational relations between the two groups.

**Table 3.** Mean values of the indicators between the non-malnutritional group (0) and the malnutritional group (1).

| Indicator | Group | Mean | Median |
| --- | --- | --- | --- |
| Left mid-upper arm circumference | 0 | 27.424 | 27.500 |
| Left mid-upper arm circumference | 1 | 24.990 | 24.200 |
| Right mid-upper arm circumference | 0 | 27.671 | 27.700 |
| Right mid-upper arm circumference | 1 | 25.110 | 24.500 |
| Left Calf Circumference | 0 | 31.627 | 31.900 |
| Left Calf Circumference | 1 | 29.228 | 29.400 |
| Right Calf Circumference | 0 | 31.667 | 31.800 |
| Right Calf Circumference | 1 | 29.270 | 29.000 |
| Left Triceps Skinfold Thickness | 0 | 19.546 | 19.000 |
| Left Triceps Skinfold Thickness | 1 | 17.368 | 18.000 |
| Right Triceps Skinfold Thickness | 0 | 19.506 | 19.000 |
| Right Triceps Skinfold Thickness | 1 | 17.244 | 17.000 |
| Left Hand Grip Strength | 0 | 14.741 | 14.200 |
| Left Hand Grip Strength | 1 | 10.425 | 10.200 |
| Right Hand Grip Strength | 0 | 15.809 | 15.200 |
| Right Hand Grip Strength | 1 | 11.764 | 12.300 |

Table 3 presents the mean and median values of each anthropometric and functional indicator for the non-malnutrition group (0) and the malnutrition group (1). Across all measured variables, systematic differences are observed between the two groups.

Overall, individuals classified as non-malnourished exhibit consistently higher central tendency values across circumference-based, skinfold-based, and functional strength measurements. Both mid-upper arm circumference and calf circumference are notably reduced in the malnutrition group. For example, the mean left mid-upper arm circumference decreases from 27.42 cm in Group 0 to 24.99 cm in Group 1, while the mean right calf circumference declines from 31.67 cm to 29.27 cm.

A similar pattern is observed for triceps skinfold thickness, with lower mean and median values in the malnutrition group, although the magnitude of difference is smaller than that observed for circumferential measurements. Functional capacity, assessed via handgrip strength, shows particularly pronounced group differences. The mean left-hand grip strength decreases from 14.74 kg in the non-malnourished group to 10.43 kg in the malnourished group, and the corresponding right-hand grip strength declines from 15.81 kg to 11.76 kg.

Collectively, these consistent reductions across structural and functional indicators in the malnutrition group provide empirical support for the discriminative validity of the GLIM classification and underscore meaningful differences in nutritional and physical status between the two populations.

### 2.3 Data imputation

Since the clinical dataset is incomplete and exhibits non-negligible missingness, especially in Day 7, multiple imputation techniques are applied to address the issue. These methods include the following:

- Last Observation Carried Forward (LOCF): Applied primarily to indicators with high missing rates, specifically body weight and BMI on day 3, use values from day 1.
- Group-wise Simple Imputation (Mean/Median): Implemented for indicators with low missing rates on day 3. Missing data within each group (malnutrition vs. normal) were imputed using the group’s mean or median values.
- Group-wise Regression Imputation: Conducted for selected indicators demonstrating high correlation with their contralateral counterparts, such as the skinfold thickness and grip strength.
- Conditional Score-based Diffusion Imputation (CSDI): An advanced deep generative method employed to handle complex data structures and severe missing data scenarios through multivariate joint imputation [21].

Imputation methods are rigorously evaluated using masking experiments, and performance metrics, including Mean Absolute Error (MAE) and Mean Squared Error (MSE), are assessed.

#### 1) LOCF Imputation (high missing rate)

We apply a Last Observation Carried Forward (LOCF) approach to Weight and BWI indicators, using day 1’s body weight/BMI to impute missing values on day 3. The result is given in Table 4.

**Table 4.** Imputation Results by LOCF Imputation.

| Indicator | MAE | MSE |
| --- | --- | --- |
| Weight 3d | 1.471 | 4.149 |
| BMI _3d | 0.590 | 0.708 |

#### 2) Group-wise Simple Imputation (low missing rate indicators, Day 3)

For all day 3 indicators except for the body weight and BMI, a group-wise masking experiment is conducted. Within each group, across the five trials, a random subset comprising 10% of the observed values is masked and imputed using the group-specific mean and median, respectively. The average MSE and MAE across all trials are listed in Table 5.

**Table 5.** Imputation results by group-wise mean or median Imputation.

| Indicator | Mean |  | Median |  |
| --- | --- | --- | --- | --- |
|  | MSE | MAE | MSE | MAE |
| Left mid-upper arm circumference_3d | 12.417 | 2.589 | 13.200 | 2.658 |
| Right mid-upper arm circumference_3d | 11.004 | 2.506 | 11.693 | 2.532 |
| Left Calf Circumference_3d | 9.580 | 2.388 | 9.814 | 2.396 |
| Right Calf Circumference_3d | 14.373 | 2.758 | 14.307 | 2.795 |
| Left Triceps Skinfold Thickness_3d | 17.629 | 3.400 | 15.249 | 3.058 |
| Right Triceps Skinfold Thickness_3d | 28.112 | 3.911 | 29.302 | 3.943 |
| Left Hand Grip Strength_3d | 33.828 | 4.500 | 33.676 | 4.503 |
| Right Hand Grip Strength_3d | 43.191 | 5.171 | 44.794 | 5.303 |

Both mean and median imputations yielded comparable performance, with mean imputation slightly outperforming median imputation for most indicators.

#### 3) Group-wise Regression Imputation (for selected indicators)

For the indicators, *left triceps skinfold thickness (3d)*, *right triceps skinfold thickness (3d)*, *left hand grip strength (3d)*, and *right-hand grip strength (3d)*, group-wise regression imputation is performed using contralateral indicators as predictors, selected based on their high correlation in the correlation matrix

Regression imputation that leverages highly correlated contralateral indicators demonstrates superior performance, notably achieving low error for skinfold thickness indicators. The results are listed in Table 6.

**Table 6.**
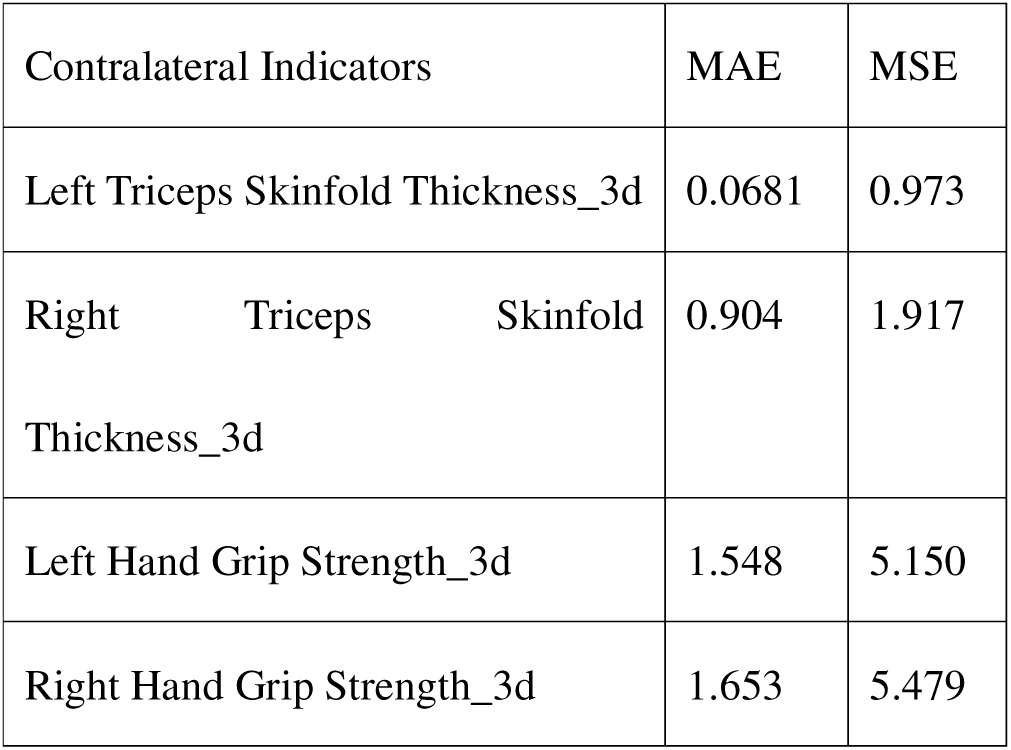
imputation results by group-wise regression imputation.

| Contralateral Indicators | MAE | MSE |
| --- | --- | --- |
| Left Triceps Skinfold Thickness_3d | 0.0681 | 0.973 |
| Right Triceps Skinfold Thickness_3d | 0.904 | 1.917 |
| Left Hand Grip Strength_3d | 1.548 | 5.150 |
| Right Hand Grip Strength_3d | 1.653 | 5.479 |

#### 4) CSDI method

The time-series dataset is split into training, validation, and test sets to train and evaluate the method. To assess the performance of the imputation method, 10% of the observed values in the validation and test sets are randomly selected and masked, serving as ground truth for evaluation. The CSDI model is trained on the training set and then used to impute the artificially masked values in the validation and test sets. For each missing value, the CSDI model generates 100 imputed samples, and the reported results represent the mean values over these 100 samples.

**Table 7.**
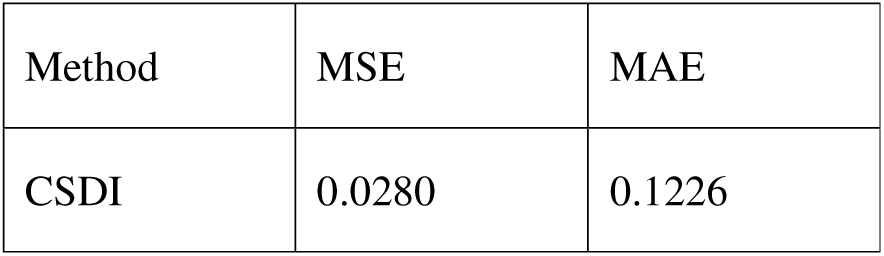
Performance of CSDI with 10% missing values.

Table 7 clearly shows that CSDI significantly outperformed the traditional methods, highlighting its robustness and suitability for addressing complex missing-data issues. *Given the superior accuracy and robustness of the CSDI method, we ultimately select it as our final imputation method* to ensure reliable and high-quality data inputs for downstream predictive modeling.

### 2.4 Indicator exploratory analysis

To guide downstream modeling tasks, we conducted a comprehensive exploratory analysis of the input indicators (indicators) using the imputed dataset. This analysis examined the distributional characteristics of the anthropometric and functional indicators across different time points, as well as their differences between malnourished and non-malnourished groups. The objective is to assess indicator behavior after imputation, evaluate temporal consistency, identify potential redundancy or complementarity among indicators, and eventually select the minimal set of indicators.

**Figure 2.**
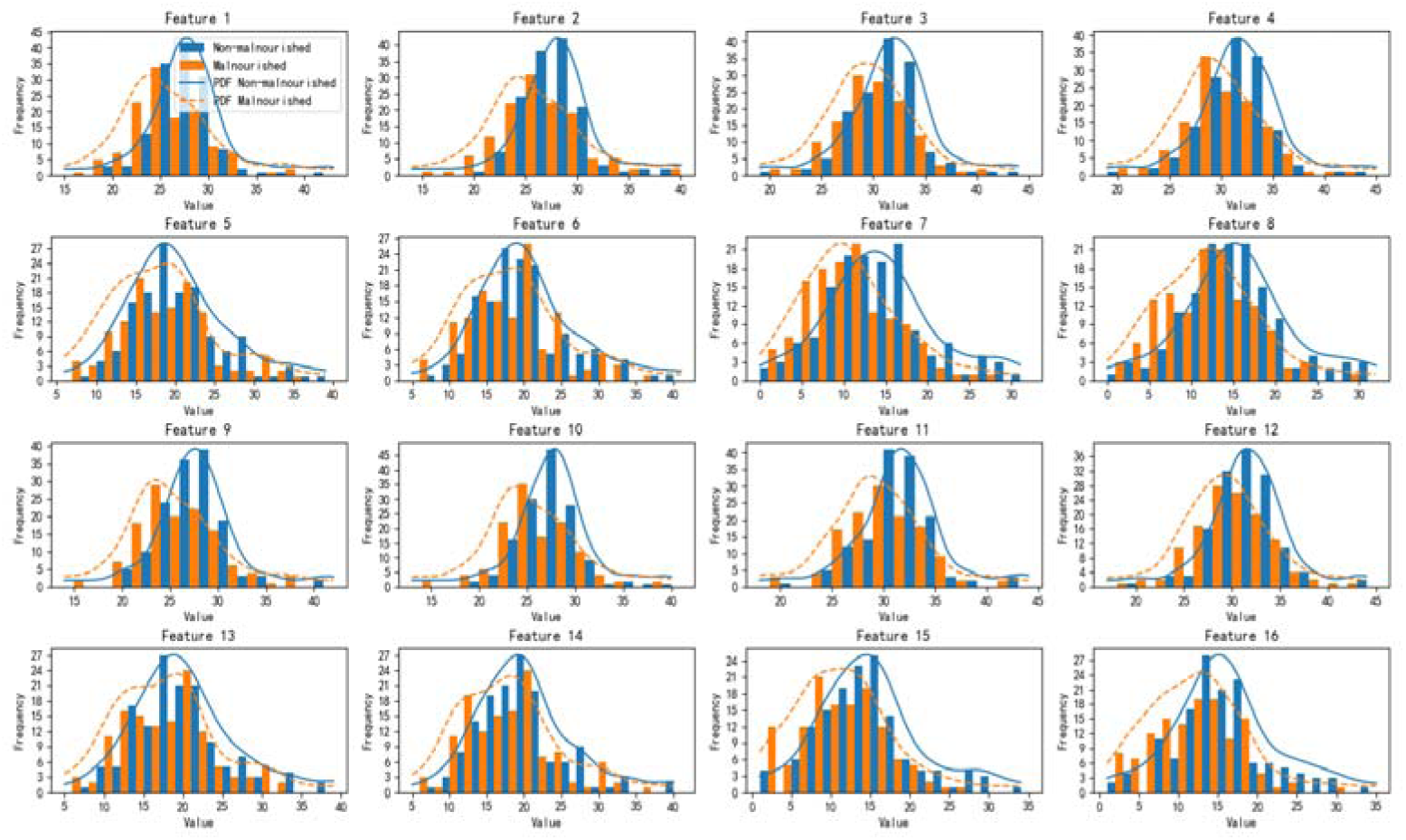
Distributions of the 16 indicators on day 1 and day 3, respectively.

#### 2.4.1 Distributions on Day 1 and Day 3

We plotted all 16 indicators (labeled F1–F16) in a 4×4 grid of histograms to visually compare their distributions between the nourished and malnourished groups on days 1 and 3, respectively. F1–F8 represent the data on day 1 of the 8 dynamic indicators, and F9–F16 represent those on day 3, respectively. We exclude the Day 7 data due to its high missingness. In each subplot, the non-malnourished group is shown with blue solid bars, and the malnourished group with orange hollow bars.

Indicators F1–F16 represent measurements collected across two time points. Specifically, F1–F8 correspond indicators MAC_L_1, MAC_R_1, CC_L_1, CC_R_1, TSF_L_1, TSF_R_1, HGS_L_1, and HGS_R_1 measured on Day 1. F9–F16 correspond to the same set of indicators measured on Day 3, denoted as MAC_L_3, MAC_R_3, CC_L_3, CC_R_3, TSF_L_3, TSF_R_3, HGS_L_3, and HGS_R_3.

To eliminate the influence of scale differences across variables, all indicators were first normalized to the range using Min-Max scaling. Consider samples. Each indicator’s mean, variance, and skewness are defined, respectively, as follows.

The composite index (CI) for each indicator j is calculated using the following formula:

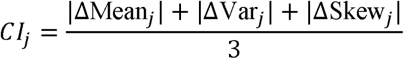

In evaluating the discriminative value of each indicator, the absolute magnitude of the between-group mean difference alone is insufficient; it must be interpreted in relation to within-group variance. As shown in the summary table, MAC (F1/F2/F9/F10) and CC (F3/F4/F11/F12) exhibit stable between-group mean differences (|ΔMean|) ranging from 0.09 to 0.11, while their corresponding within-group variances (Var□/Var□) remain as low as 0.01–0.02 — the lowest across all indicators. This indicates that both MAC and CC achieve meaningful between-group separation with minimal within-group dispersion. HGS (F7/F8/F15/F16), despite showing slightly larger mean differences (0.11–0.13), exhibits within-group variances of 0.03–0.04, two to four times those of MAC/CC. The between-group shift in HGS therefore occurs against a backdrop of considerable within-group variability, resulting in blurred decision boundaries and substantial dilution of discriminative information. TSF (F5/F6/F13/F14) fares even less favorably: its mean differences are the smallest of all four indicator types (0.06–0.07), while its variance remains comparably elevated at 0.03, placing it at a disadvantage on both dimensions simultaneously. These quantitative findings are fully consistent with the visual evidence from the histograms — MAC and CC display sharp, well-separated KDE peaks, whereas TSF and HGS show broad, flat curves with substantial distributional overlap between groups.

**Table 8:**
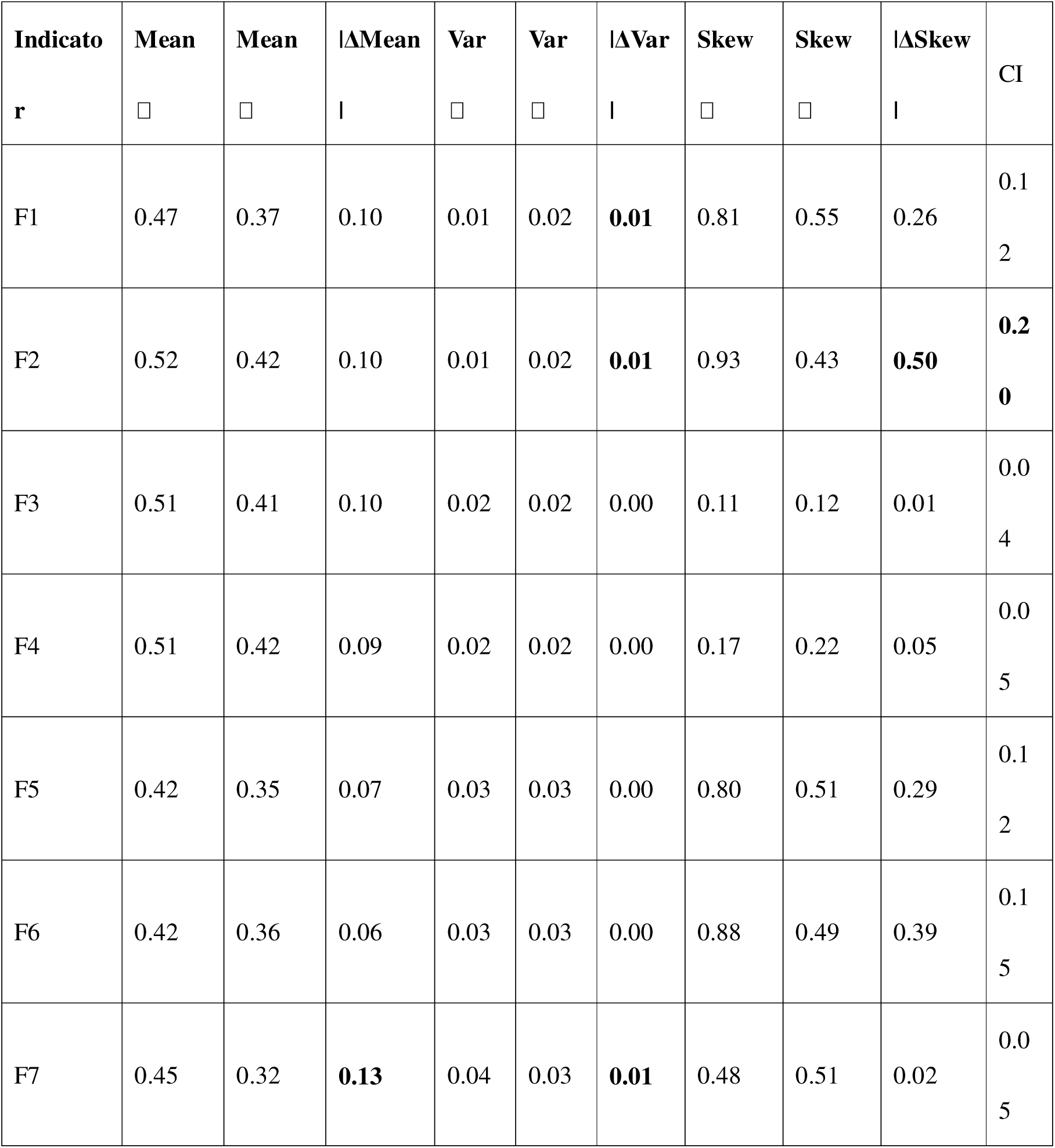
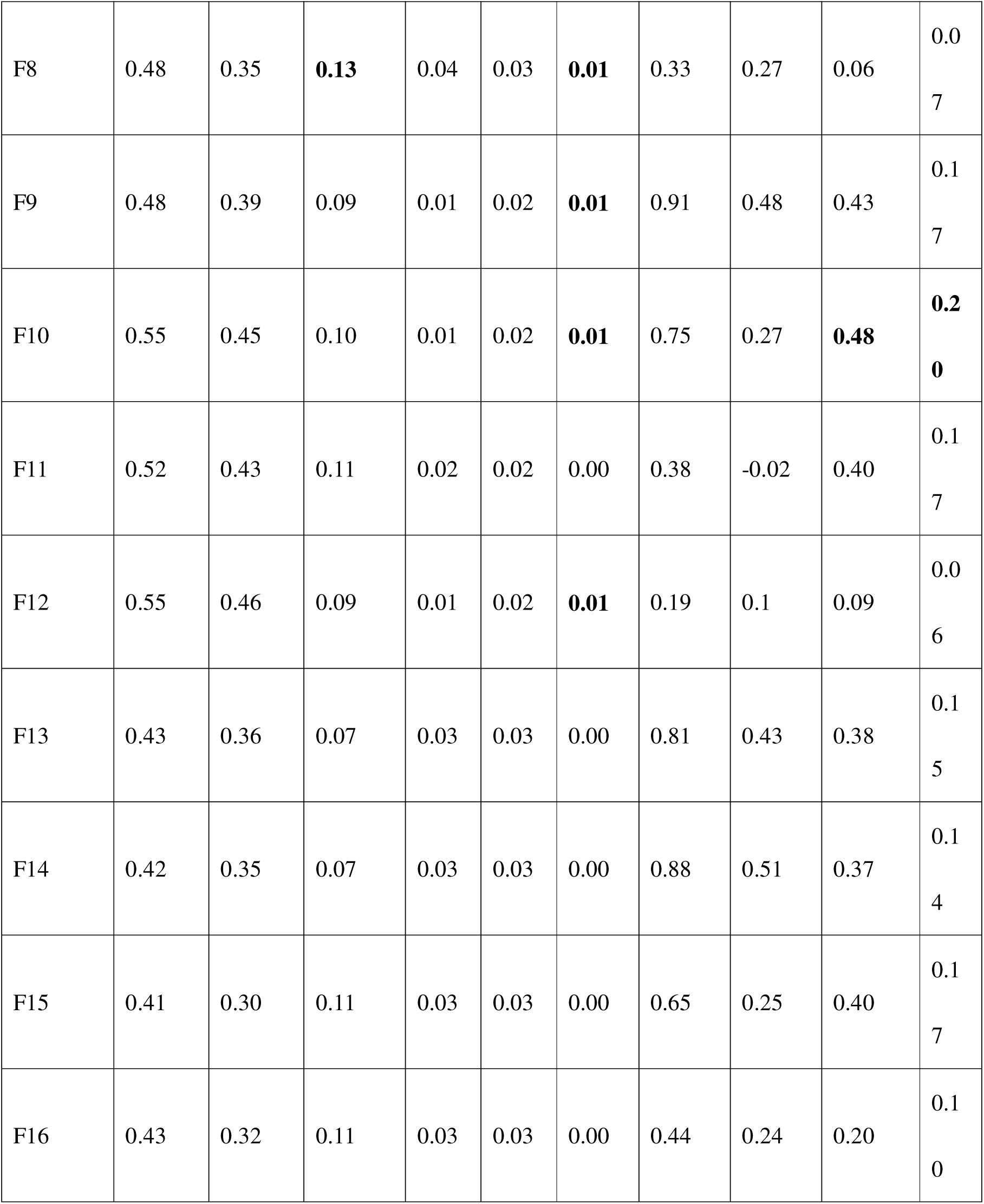
Summary of group-wise distribution means, variances, and skewness, their differences between Day 1 and Day 3, and the Composite Index (CI) for each indicator. The two highest values in each difference between the two days are highlighted in boldface.

| Indicato<br>r | Mean<br>□ | Mean<br>□ | ΔMean<br>\| | Var<br>□ | Var<br>□ | ΔVar<br>\| | Skew<br>□ | Skew<br>□ | ΔSkew<br>\| | CI |
| --- | --- | --- | --- | --- | --- | --- | --- | --- | --- | --- |
| F1 | 0.47 | 0.37 | 0.10 | 0.01 | 0.02 | **0.01** | 0.81 | 0.55 | 0.26 | 0.1<br>2 |
| F2 | 0.52 | 0.42 | 0.10 | 0.01 | 0.02 | **0.01** | 0.93 | 0.43 | **0.50** | **0.2**<br>**0** |
| F3 | 0.51 | 0.41 | 0.10 | 0.02 | 0.02 | 0.00 | 0.11 | 0.12 | 0.01 | 0.0<br>4 |
| F4 | 0.51 | 0.42 | 0.09 | 0.02 | 0.02 | 0.00 | 0.17 | 0.22 | 0.05 | 0.0<br>5 |
| F5 | 0.42 | 0.35 | 0.07 | 0.03 | 0.03 | 0.00 | 0.80 | 0.51 | 0.29 | 0.1<br>2 |
| F6 | 0.42 | 0.36 | 0.06 | 0.03 | 0.03 | 0.00 | 0.88 | 0.49 | 0.39 | 0.1<br>5 |
| F7 | 0.45 | 0.32 | **0.13** | 0.04 | 0.03 | **0.01** | 0.48 | 0.51 | 0.02 | 0.0<br>5 |
| F8 | 0.48 | 0.35 | **0.13** | 0.04 | 0.03 | **0.01** | 0.33 | 0.27 | 0.06 | 0.07 |
| F9 | 0.48 | 0.39 | 0.09 | 0.01 | 0.02 | **0.01** | 0.91 | 0.48 | 0.43 | 0.17 |
| F10 | 0.55 | 0.45 | 0.10 | 0.01 | 0.02 | **0.01** | 0.75 | 0.27 | **0.48** | **0.20** |
| F11 | 0.52 | 0.43 | 0.11 | 0.02 | 0.02 | 0.00 | 0.38 | -0.02 | 0.40 | 0.17 |
| F12 | 0.55 | 0.46 | 0.09 | 0.01 | 0.02 | **0.01** | 0.19 | 0.1 | 0.09 | 0.06 |
| F13 | 0.43 | 0.36 | 0.07 | 0.03 | 0.03 | 0.00 | 0.81 | 0.43 | 0.38 | 0.15 |
| F14 | 0.42 | 0.35 | 0.07 | 0.03 | 0.03 | 0.00 | 0.88 | 0.51 | 0.37 | 0.14 |
| F15 | 0.41 | 0.30 | 0.11 | 0.03 | 0.03 | 0.00 | 0.65 | 0.25 | 0.40 | 0.17 |
| F16 | 0.43 | 0.32 | 0.11 | 0.03 | 0.03 | 0.00 | 0.44 | 0.24 | 0.20 | 0.10 |

Low within-group variance not only signals strong discriminative power but also reflects the intrinsic measurement stability of an indicator. The consistently low within-group variance in MAC and CC indicates that individuals with the same nutritional status yield highly homogeneous measurements on these indicators, suggesting strong clinical reproducibility and discriminative purity. This stability is further corroborated in the longitudinal dimension: comparing Day 1 indicators (F1–F4) with their Day 3 counterparts (F9–F12), both MAC and CC maintain highly consistent distributional shapes and stable between-group difference patterns across time points, demonstrating good reproducibility and robustness to short-term physiological fluctuation.

At the level of distributional shape, the absolute skewness difference (|ΔSkew|) provides a layer of discriminative information beyond mean and variance, capturing whether the two groups exhibit systematic divergence in the asymmetry of their distributions. For MAC, all four indicators (F1/F2/F9/F10) yield |ΔSkew| values ranging from 0.26 to 0.50, with F2 (MAC_R_1) and F10 (MAC_R_3) reaching 0.50 and 0.48, respectively — the highest values among all 16 indicators. This consistent pattern indicates that MAC not only separates the two groups in terms of central tendency but also reveals systematic structural divergence in distributional shape, reflecting fundamental biological differences in body composition between malnourished and non-malnourished individuals. CC, however, shows uneven performance on this dimension: F11 (CC_L_3) achieves a |ΔSkew| of 0.40, but the remaining three CC indicators (F3/F4/F12) yield values of only 0.01–0.09, indicating insufficient internal consistency. The case for CC on this dimension is therefore only partially supported, and its prioritization rests more firmly on the first two dimensions — group means difference and within-group variance, and longitudinal stability. In contrast, HGS indicators F7 and F8 yield |ΔSkew| values of merely 0.02 and 0.06, approaching zero, indicating that the two groups are virtually indistinguishable in grip strength distributional shape and offer no meaningful structural discriminative signal. TSF shows intermediate |ΔSkew| values (0.29–0.39), but given its concurrent disadvantages of high variance and low mean difference, this degree of skewness divergence is unlikely to translate into practical discriminative capacity.

Taking into account all three metrics, MAC demonstrates comprehensive superiority in combined performance across between-group mean difference, within-group variance, measurement stability and longitudinal reproducibility, and skewness structure; CC performs comparably well on the first two dimensions. Jointly, these two indicators provide robust statistical justification for their prioritization, while the systematic disadvantages of TSF and HGS across multiple dimensions further reinforce this conclusion. The superior discriminative power of MCC and the strong performance of CC are also evident in the composite index (CI). This evidence strongly supports our selection of MCC and CC as the essential metrics for assessing malnutrition risk using longitudinal patient data.

## 3 Methods

We employ a host of machine learning methods to perform the intended classification task and package them into a clinically usable toolbox.

### 3.1 Machine learning methods

We briefly review the machine learning algorithms used for classification tasks in this study.

- <u>Logistic Regression</u>: Logistic regression [22] is a simple, yet robust classification algorithm. It uses a sigmoid function to convert a linear combination of input indicators into binary outputs (0 or 1) for binary classification. In the model, unscaled indicators with large magnitudes can bias the weights and affect convergence during training.
- <u>SVM</u>: Support Vector Machines (SVMs) [23] seek the “hyperplane” in indicator space that maximizes the margin between classes. When the data are not linearly separable, the “kernel trick” projects them into a higher-dimensional space where a linear separator may exist. SVMs rely on dot products and distance computations; unscaled indicators may dominate and distort the margin.
- <u>KNN</u>: k-Nearest neighbors (KNN) [24] is a simple yet effective classification algorithm. It assigns labels to new inputs based on their proximity to training samples with known labels, using majority voting or other distance-based methods. KNN is highly sensitive to indicator scaling because distance-based voting tends to dominate.
- <u>Decision tree</u>: A decision tree [25] is a non-parametric classifier that recursively partitions the indicator space: at each internal node, it picks the best indicator and threshold that splits the incoming samples into two child nodes of higher “purity,” and it continues splitting until some stopping criterion is met (e.g., maximum depth, minimum samples per node, etc.). Decision trees are invariant to indicator scaling because they only use comparisons of indicator values (e.g., “is indicator > threshold”).
- <u>Random Forest</u>: Random forest [26] is a bagging-based ensemble learning method that combines multiple decision trees to create a strong classifier. It randomly selects a subset of indicators and samples from the training data to build each tree, resulting in a robust and efficient model. Like single decision trees, random forests split based on indicator thresholds, making them robust for unscaled data.
- <u>XGBOOST</u>: XGBoost [27] is a gradient boosting-based ensemble learning method that builds decision trees sequentially, where each new tree focuses on correcting the errors of the previous ones. As a tree-based model, XGBoost uses threshold-based splitting, making it insensitive to the scale of the input indicators.

## Ensemble methods

To further enhance predictive performance and robustness, we construct an ensemble framework that integrates multiple base classifiers with heterogeneous modeling characteristics. The base learners are designed to capture complementary decision patterns, ranging from linear structures to complex nonlinear relationships.

For each sample *x*, suppose we have *K* trained base models. Each model produces a probability estimate of the positive class:

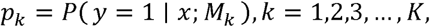

where *M*_k_denotes the *k*-th classifier. The predicted probabilities are then concatenated to form a meta-indicator vector

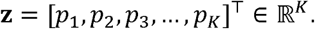

Rather than directly averaging these outputs, we train an additional Logistic Regression model on the training set using z as input indicators to learn an optimal combination of the base predictions. The final ensemble prediction is computed as follows

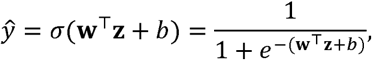

where **w** and *b* are learnable parameters and a(,)denotes the sigmoid function.

This ensemble strategy enables the model to adaptively weight the contributions of different classifiers, thereby improving overall stability and generalization performance.

## SHAP analysis

SHAP (Shapley Additive Explanations), introduced by Lundberg and Lee [28], was applied to the three-time-point XGBoost model and the best-performing ensemble configurations, specifically the two-time-point ensemble. The analysis was conducted to quantify the contribution of individual indicators to the prediction of malnutrition risk (Class 1). Across these models, the SHAP results revealed consistent, clinically coherent attribution patterns, indicating that a limited subset of dynamic and static indicators primarily drove the models’ predictions.

## 4. Results

We train the models using the preprocessed dataset of 269 patients following the machine learning protocol with the split ratio among the training, validation, and test datasets at 7:1:2. In the following, we evaluate model performance under different temporal and indicator configurations and then conduct SHAP analyses for the best-performing models: the three-time-point XGBoost model and the top ensemble model.

### 4.1 Performance assessment

We evaluate model performance under a structured experimental design that jointly considers temporal and indicator configurations. From a temporal perspective, four settings are examined: (1) Day 1 only, (2) Day 7 only, (3) Day 1 + Day 3, and (4) Day 1 + Day 3 + Day 7. For each temporal configuration, two indicator configurations are evaluated: (i) static variables combined with the full set of dynamic indicators, and (ii) static variables combined with selected dynamic indicators (MAC and CC only). This factorial design enables a systematic comparison of how longitudinal depth and indicator richness interact to influence classification performance. Model performance across all configurational combinations is assessed using accuracy, precision, recall, F1 score, AUC–ROC, and MCC to identify the optimal temporal–indicator pairing for downstream prediction tasks.

Model’s hyperparameters are selected based on performance on the validation set. After identifying the “optimal hyperparameters”, the models are retrained on the combined training and validation sets, and the final performance is evaluated on the held-out test set to report the assessment metrics.

#### 4.1.2 Performance using indicators from Day 1

When all dynamic indicators were included (Table 9), most models achieved accuracy values between 0.76 and 0.80. The ensemble model yielded the highest accuracy (0.8148) and F1 score (0.7917), while KNN achieved the highest AUROC (0.8063). XGBoost achieved the highest recall (0.8462), indicating greater sensitivity to the positive class, although its overall AUROC was comparable to those of other models. Overall, performance across models was relatively stable, with moderate variation among different classifiers.

**Table 9.**
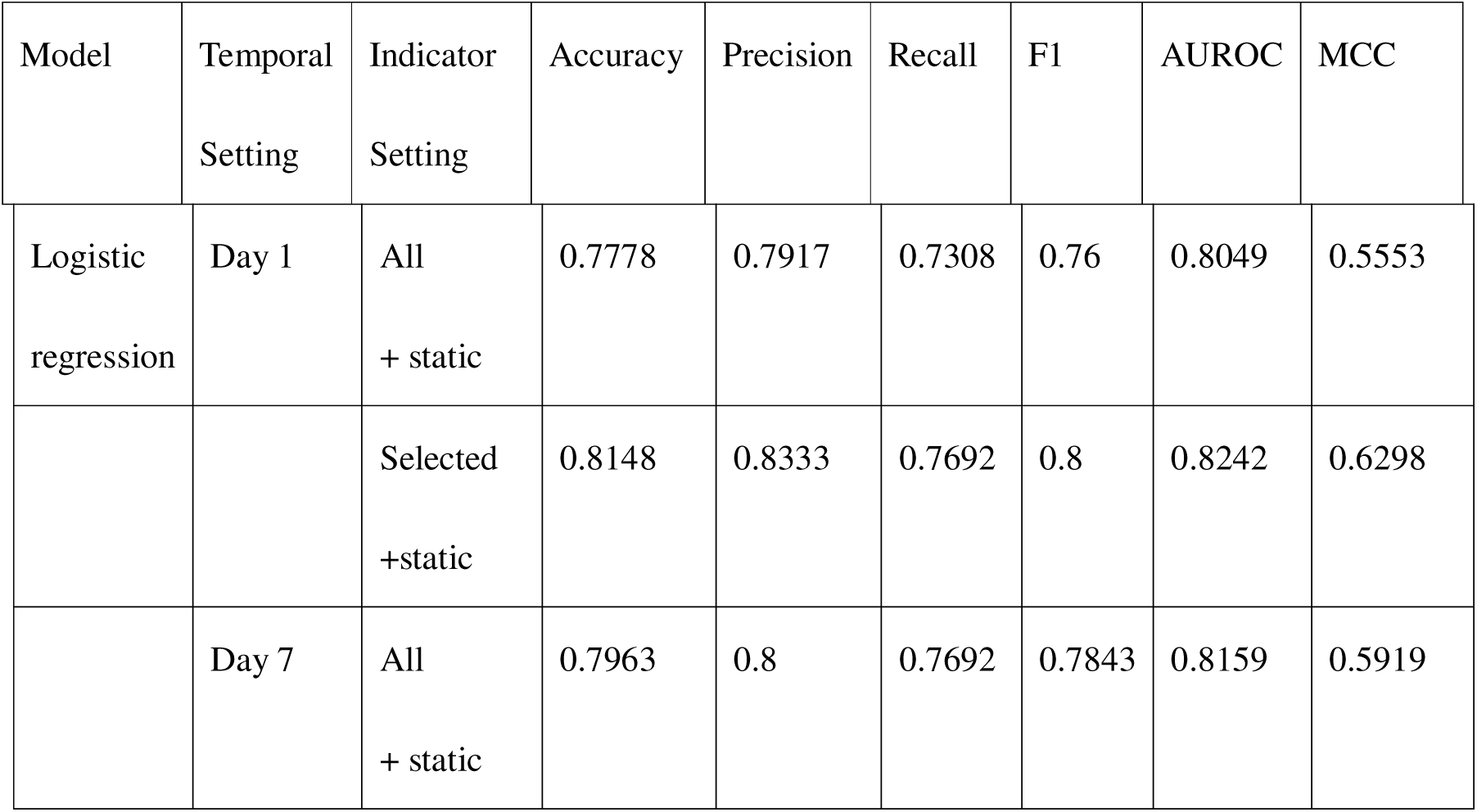
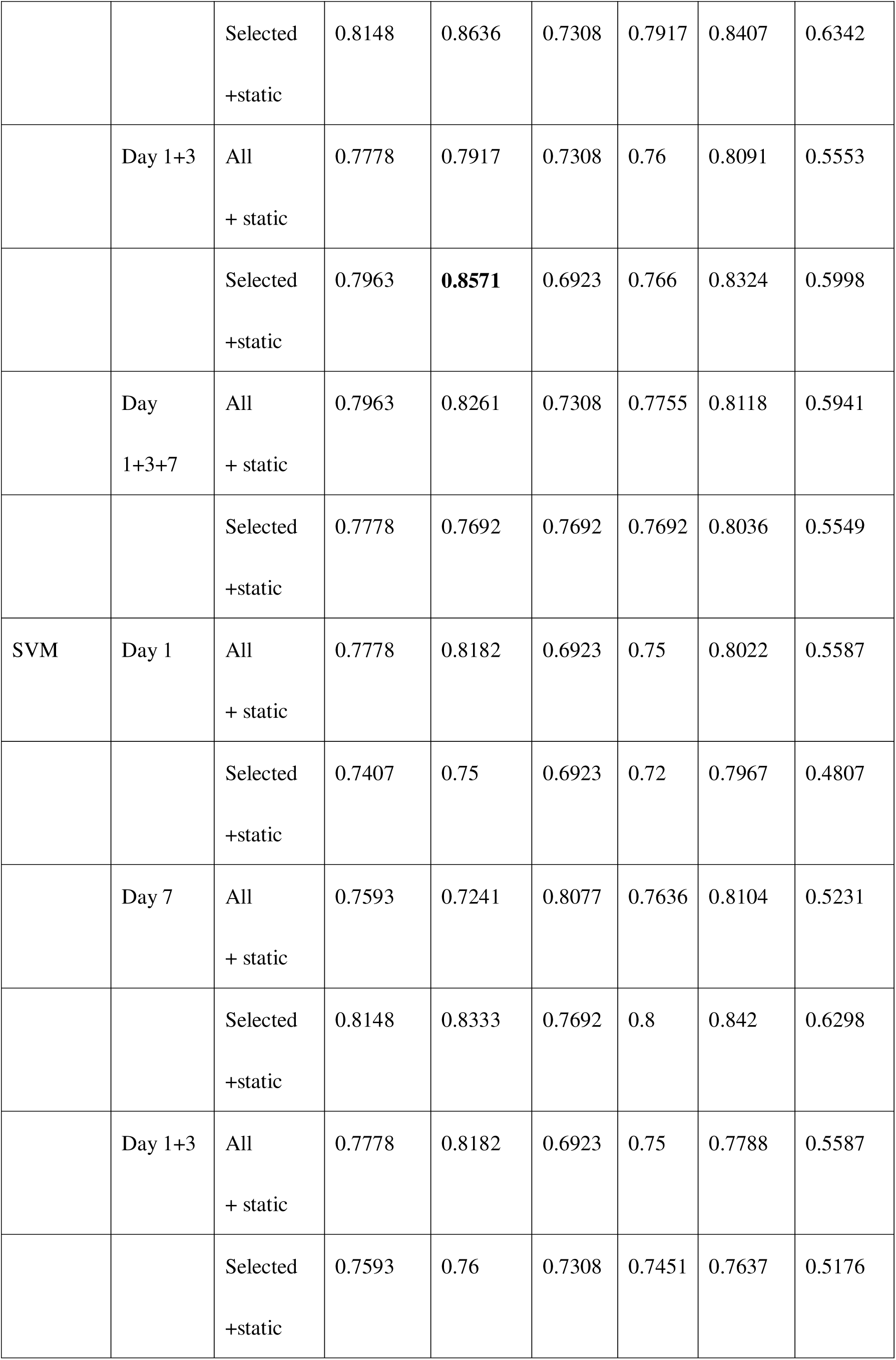
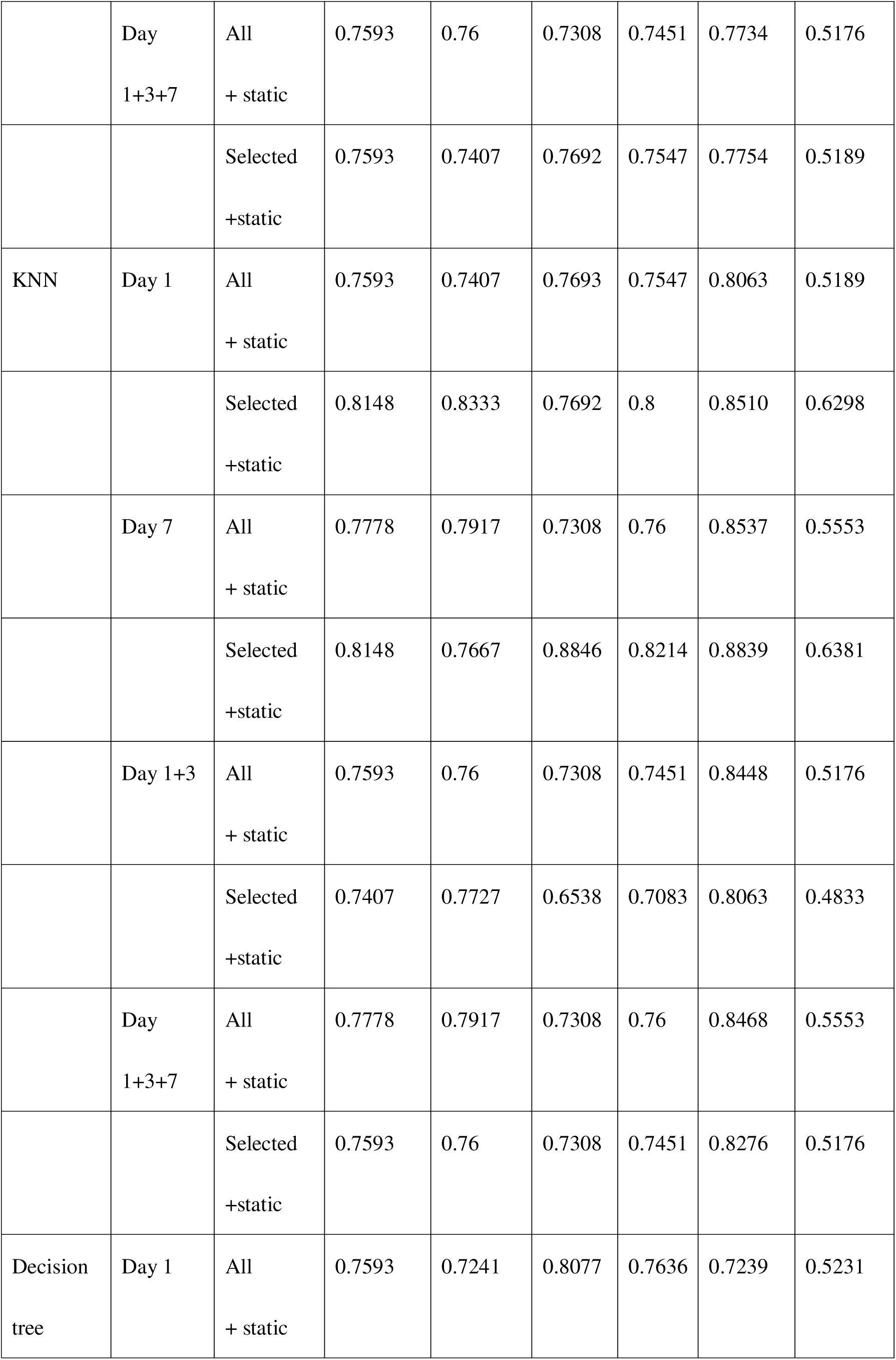
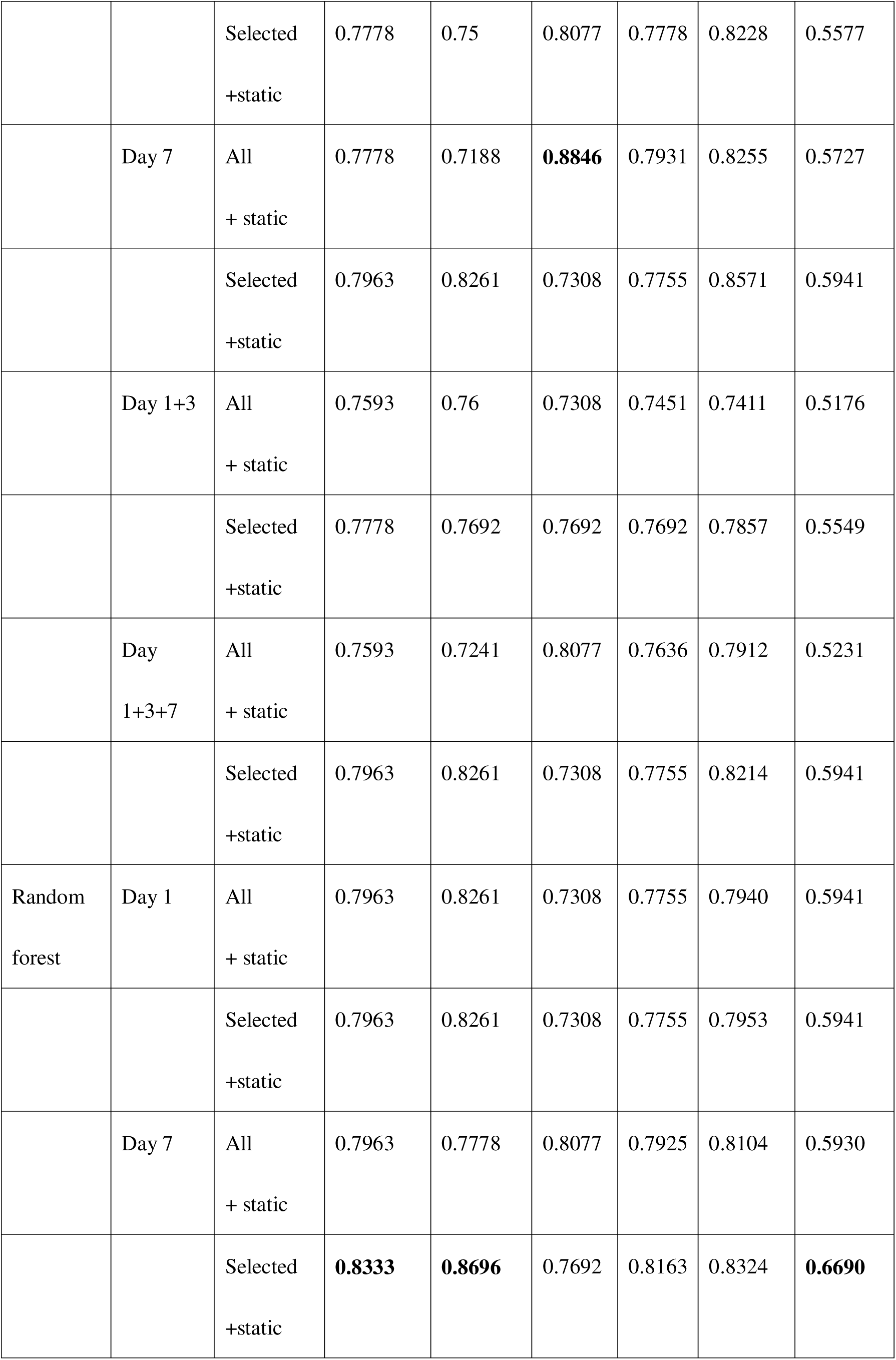
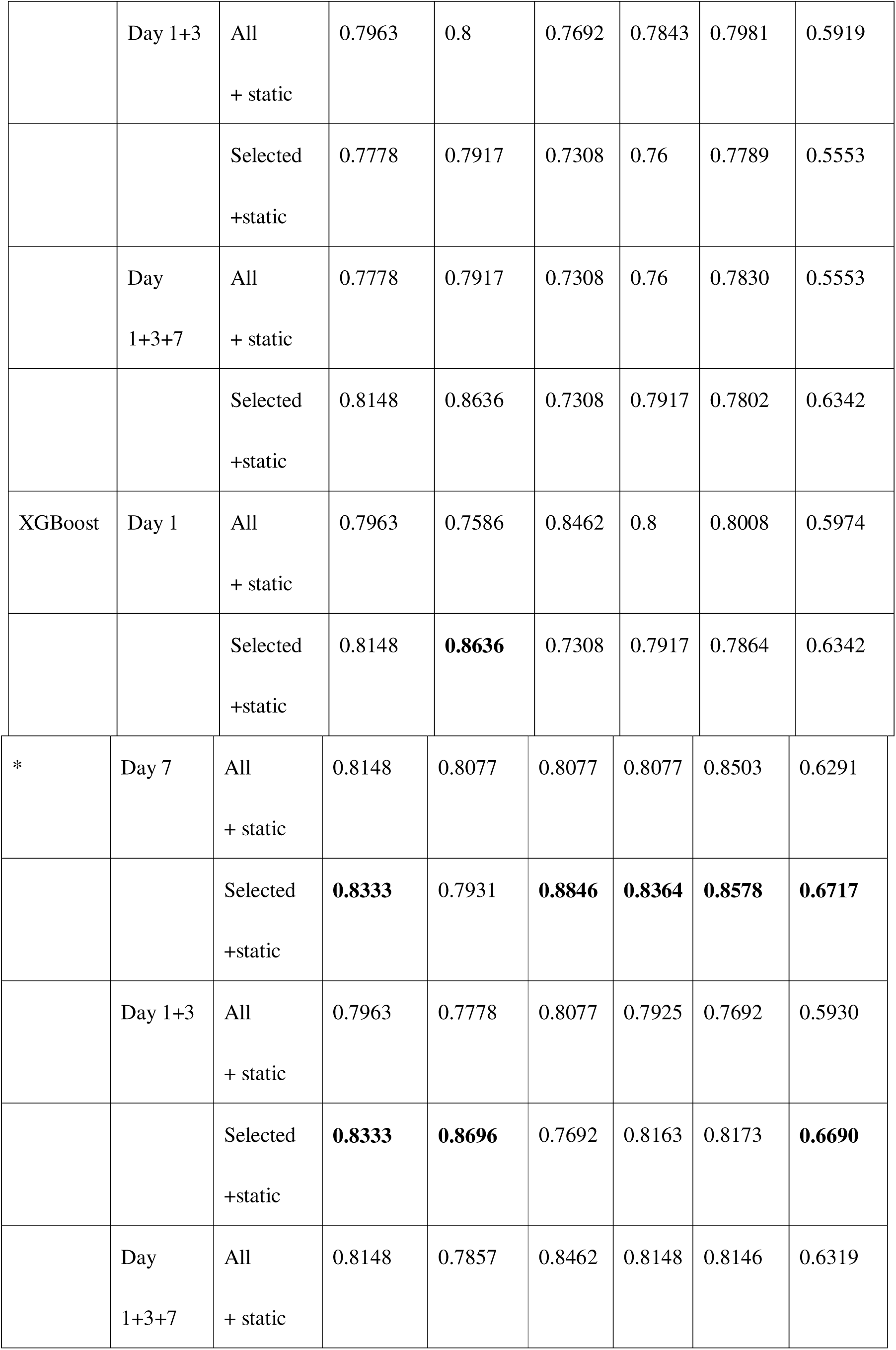
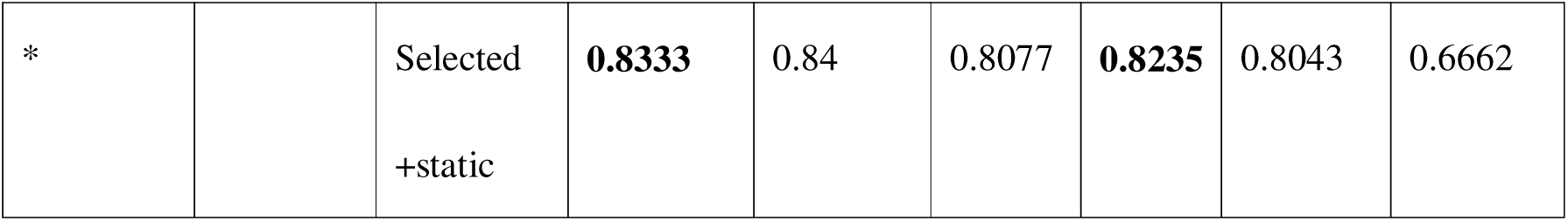
Performance of individual models across temporal and indicator configurations.

In contrast, when only selected dynamic indicators (MAC and CC) were combined with static variables, performance did not deteriorate; in several cases, it improved. The ensemble model achieved the highest overall performance, with an accuracy of 0.8333, an F1 score of 0.8235, and an AUROC of 0.8503. Notably, both Logistic Regression and KNN achieved higher AUROC values (0.8242 and 0.8510, respectively) than their counterparts using the full indicator set. These results suggest that the reduced indicator configuration retains substantial discriminative power while potentially mitigating redundancy.

A direct comparison in the table indicates that incorporating all dynamic variables does not necessarily yield superior classification performance on Day 1. The selected indicator configuration achieves comparable or improved performance across several models, particularly for AUROC and F1 score. This finding implies that MAC and CC measurements, when combined with static characteristics, capture a substantial proportion of the predictive signal within the full dynamic indicator set.

#### 4.1.3 Performance using indicators from Day 7

Under the full dynamic indicator setting, KNN achieves the highest AUROC (0.8537) among all individual models, indicating strong ranking capability despite moderate accuracy. XGBoost follows closely with an AUROC of 0.8503, while maintaining more balanced precision and recall. Logistic Regression and Random Forest exhibit comparable overall performance, both reaching an accuracy of 0.7963 with stable discrimination (AUROC ≈ 0.81). The Decision Tree yields the highest recall (0.8846), indicating strong sensitivity to the positive class, although its precision is relatively low. The ensemble model slightly outperforms the individual classifiers, but the relative ordering among the individual models remains consistent.

When restricting the indicator space to selected dynamic variables (MAC and CC), Logistic Regression and SVM both reach an accuracy of 0.8148, demonstrating improved stability under reduced dimensionality. KNN and XGBoost show notably high recall values (0.8846), indicating enhanced detection of malnourished cases. Random Forest achieves the highest precision (0.8696), reflecting conservative but reliable classification behavior. Although the ensemble model achieves the best overall metrics, several individual classifiers achieve similar performance, suggesting that the discriminative signal on Day 7 is sufficiently strong to be captured even without model aggregation.

Despite their superior performance to models trained on Day 1 data, we reserve judgment on these models due to substantial data missingness.

#### 4.1.4 Performance using indicators from Day 1 and Day 3

We built classification models using all available data from day 1 and day 3. Table 9 reports the results when combining measurements from Day 1 and Day 3 under the two indicator configurations.

Using the complete set of dynamic indicators, model performance remains moderate, with accuracy ranging from 0.7593 to 0.7963. Among individual classifiers, KNN achieves the highest AUROC (0.8448) and XGBoost the highest recall (0.8077). Among ensemble models, Log+DT+XGB achieves the highest AUROC (0.8187), while RF+XGB matches the highest accuracy observed among individual classifiers (0.7963).

When restricting the indicator space to selected dynamic variables, performance becomes more differentiated. XGBoost achieves the highest accuracy (0.8333) and F1 score (0.8163) among individual classifiers, while Logistic Regression maintains strong precision (0.8571) and stable discrimination (AUROC = 0.8324). The Log+DT+XGB ensemble achieves the strongest overall performance across all models, with an accuracy of 0.8519 and AUROC of 0.8352.

Comparing the two indicator configurations, the selected dynamic indicator setting does not reduce predictive capability and, in some cases, improves overall performance, particularly for boosting-based models. This suggests that including additional dynamic variables beyond MAC and CC does not necessarily yield incremental benefit when early time-point data are accounted for.

#### 4.1.5 Performance using raw indicators from 3 days

The joint incorporation of measurements from Day 1, Day 3, and Day 7 provides a comprehensive longitudinal representation of each patient. Table 9 presents the resulting classification performance under the two indicator configurations.

With the complete dynamic indicator set, model performance remains stable but does not markedly surpass the best results observed under the Day 1 + Day 3 configuration (Section 4.1.4). Among individual classifiers, XGBoost achieves the highest F1 score (0.8148) and strong recall (0.8462), indicating effective sensitivity when multi-time-point information is used. KNN achieves the highest AUROC (0.8468), indicating strong ranking performance in the expanded temporal indicator space. In contrast, Logistic Regression and Random Forest exhibit consistent but moderate performance, with accuracy values remaining below 0.80.

Under the reduced dynamic indicator configuration, XGBoost continues to demonstrate strong overall performance (accuracy = 0.8333, F1 = 0.8235), while Random Forest achieves the highest precision (0.8636), suggesting reliable positive-class identification. Although ensemble approaches demonstrate stable metrics across settings, their advantage over high-performing single models is limited under this configuration.

A comparison with the two-time-point configuration (Day 1 + Day 3) indicates that incorporating all three time points does not result in substantial additional improvement. While expanding from a single to two time points enhances classification stability, the transition from two to three time points yields only marginal performance gains. This suggests that most of the longitudinal discriminative information may already be captured when combining the first two time points, and the incremental contribution of the third time point appears limited.

Among the evaluation metrics, AUROC exhibits the clearest temporal pattern. Day 7 configurations generally yield higher AUROC values than Day 1, indicating stronger class separation at the later stage. This trend is particularly evident under the selected dynamic configuration, where KNN achieves the highest AUROC (0.8839), and XGBoost also demonstrates strong ranking performance (AUROC = 0.8578). Notably, the selected dynamic setting frequently matches or exceeds the full dynamic configuration in AUROC across temporal combinations.

With respect to recall, tree-based models demonstrate stronger sensitivity overall. Under the Day 7 configuration, recall above 0.88 is achieved by Decision Tree under the full indicator setting and by XGBoost under the selected indicator setting, suggesting that high sensitivity at this time point is attainable but model– and configuration-dependent. In certain temporal settings, especially on Day 1 and Day 3, the full dynamic indicator setting shows a mild advantage in recall, though this tendency is not consistently observed across all models.

From an indicator-configuration perspective, the selected dynamic variables (MAC and CC) generally perform comparably to or better than the full dynamic indicator set, suggesting that core anthropometric indicators capture the predominant predictive signal without requiring the full set of variables.

Overall, the results indicate that later-time-point information strengthens class discrimination, that tree-based models exhibit heightened sensitivity, and that core dynamic indicators are sufficient to support classification. The incremental benefit of adding a third time point beyond Day 1 + Day 3 is minimal and inconsistent, offering no substantial additional discriminative value.

**Table 10.** Cross-configuration performance comparison of best-performing ensemble models under All+Static and Selected+Static indicator settings.

| Temp<br>oral<br>Settin<br>g | Indicator<br>Setting | Ensemble<br>configuration | Accuracy | Precisio<br>n | Recall | F1 | AURO<br>C | MCC |
| --- | --- | --- | --- | --- | --- | --- | --- | --- |
| Day 1 | All + static | Rf+xgb | 0.8148 | 0.8636 | 0.7308 | 0.7917 | 0.8077 | 0.6342 |
|  |  | Knn+dt+xgb | 0.7963 | 0.7586 | 0.8462 | 0.8 | 0.7995 | 0.5974 |
|  | Selected+sta<br>tic | Rf+xgb | 0.8148 | 0.8333 | 0.7692 | 0.8 | 0.7926 | 0.6298 |
| * |  | Knn+dt+xgb | 0.8333 | 0.84 | 0.8077 | 0.8235 | 0.8503 | 0.6662 |
| Day 7 | All + static | Dt+xgb | 0.8148 | 0.7667 | **0.8846** | 0.8214 | **0.8695** | 0.6381 |
|  |  | Rf+xgb | 0.8148 | 0.8333 | 0.7692 | 0.8 | 0.8503 | 0.6298 |
| * | Selected+sta<br>tic | Dt+xgb | 0.8333 | 0.84 | 0.8077 | 0.8235 | 0.8716 | 0.6662 |
|  |  | Rf+xgb | 0.8333 | 0.7742 | 0.9231 | 0.8421 | 0.8585 | 0.6801 |
| Day<br>1+3 | All + static | Rf+xgb | 0.7963 | 0.8261 | 0.7308 | 0.7755 | 0.8036 | 0.5941 |
|  |  | Log+dt+xgb | 0.7778 | 0.75 | 0.8077 | 0.7778 | 0.8187 | 0.5577 |
|  | Selected+static | Rf+xgb | 0.8148 | 0.8636 | 0.7308 | 0.7917 | 0.8132 | 0.6342 |
| * |  | Log+dt+xgb | **0.8519** | **0.875** | 0.8077 | **0.84** | 0.8352 | **0.7044** |
| Day<br>1+3+7 | All + static | Log+xgb | 0.8148 | 0.7857 | 0.8462 | 0.8148 | 0.8407 | 0.6319 |
|  |  | Rf+xgb | 0.7963 | 0.7777 | 0.8077 | 0.7925 | 0.8104 | 0.5930 |
| * | Selected+static | Log+xgb | 0.8333 | 0.84 | 0.8077 | 0.8235 | 0.8063 | 0.6662 |
|  |  | Rf+xgb | 0.8148 | 0.8333 | 0.7692 | 0.8 | 0.8077 | 0.6298 |

The summarized ensemble results reveal clear performance patterns across temporal and indicator configurations.

Across different temporal and indicator configurations, the ensemble models demonstrate consistently stable performance, with accuracy ranging from 0.77 to 0.85 and AUROC values generally exceeding 0.80, reflecting the fundamental robustness of the constructed predictive framework.

From a temporal perspective, Day 7 consistently achieves the strongest discriminative performance across all evaluated configurations. Under the All+Static setting, the DT+XGB ensemble reaches an AUROC of 0.8695; under the Selected+Static setting, the same model further improves to an AUROC of 0.8716, the highest value observed across all ensemble results. Particularly noteworthy is the RF+XGB model under the Day 7 Selected configuration, which attains the highest recall (0.9231) and F1 score (0.8421), demonstrating outstanding sensitivity in identifying high-risk patients. These results collectively corroborate the importance of Day 7 as a critical discriminative time point, likely reflecting the progressive physiological divergence between malnourished and non-malnourished patients as hospitalization advances, thereby rendering later-stage measurements increasingly powerful for distinguishing between groups.

Regarding indicator configuration, a systematic comparison of the same ensemble model under All and Selected indicator settings reveals that the Selected configuration confers clear advantages in most cases. At Day 1, the KNN+DT+XGB model improves its MCC from 0.5974 to 0.6662 (+0.069) and AUROC from 0.7995 to 0.8503 under the Selected setting. At Day 7, RF+XGB achieves an MCC gain of +0.050, rising from 0.6298 to 0.6801. At Day 1+3, Log+DT+XGB exhibits the largest improvement across the entire study, with accuracy increasing from 0.7778 to 0.8519 and MCC from 0.5577 to 0.7044 (+0.147), representing the highest MCC and accuracy observed in any configuration. At Day 1+3+7, both accuracy and MCC improve under the Selected setting, although AUROC declines slightly, suggesting that indicator selection enhances the classification boundary under multi-time-point integration but exerts a modest adverse effect on probability ranking. Across all eight same-model comparisons, seven satisfy Selected ≥ All in terms of MCC, indicating that the selected indicator set—comprising bilateral mid-upper arm circumference and calf circumference—already captures the predominant discriminative signal, and that incorporating the full dynamic variable set does not systematically enhance ensemble model performance.

Regarding model combination, ensemble architectures incorporating XGBoost (RF+XGB, DT+XGB, Log+XGB) consistently outperform across configurations, underscoring the central role of gradient boosting within the ensemble framework. In contrast, three-component combinations such as KNN+DT+XGB, while stable, do not substantially outperform structurally simpler two-component ensembles, suggesting that increasing model complexity does not yield proportionate performance gains in small-sample clinical settings. Overall, the selected anthropometric indicators demonstrate sufficient expressive capacity, while XGBoost-based ensemble models exhibit strong robustness and predictive efficiency across diverse temporal and indicator configurations, establishing them as the most representative high-performance modeling solution within the proposed framework.

### 4.2 SHAP analysis

SHAP (Shapley Additive Explanations) analysis was applied to the ensemble model (Log+DT+XGB) trained on the minimal dataset from Day 1 and Day 3 and to the XGBoost model trained on all three time points under the selected indicator configuration, to quantify the contribution of individual indicators to malnutrition risk prediction (Class 1). All SHAP summary bar plots and beeswarm plots were generated using the top 10 most influential indicators in each configuration.

For the ensemble model (Log+DT+XGB) trained on the dataset from the first two time points (Day 1 and Day 3), age group emerges as the primary driver, with the highest mean absolute SHAP value, followed by early anthropometric indicators MAC_L1, MAC_R3, CC_L1, MAC_R1, and CC_R3. Static factors, such as injury location, contribute to a comparatively lesser degree. The beeswarm plot reveals a generally consistent directional pattern: higher age-group values are predominantly associated with positive SHAP values, indicating an increased predicted risk of malnutrition. For anthropometric indicators, lower MAC and CC values tend to cluster in the positive SHAP region, reflecting increased risk, while higher values are associated with negative SHAP contributions. However, the directional patterns are relatively dispersed, with SHAP values ranging from approximately −0.04 to +0.04, reflecting a moderate, somewhat heterogeneous discriminative signal available at earlier stages of hospitalization. Nevertheless, the age group retains robust explanatory power, underscoring its role as a stable baseline predictor.

**Figure 3.**
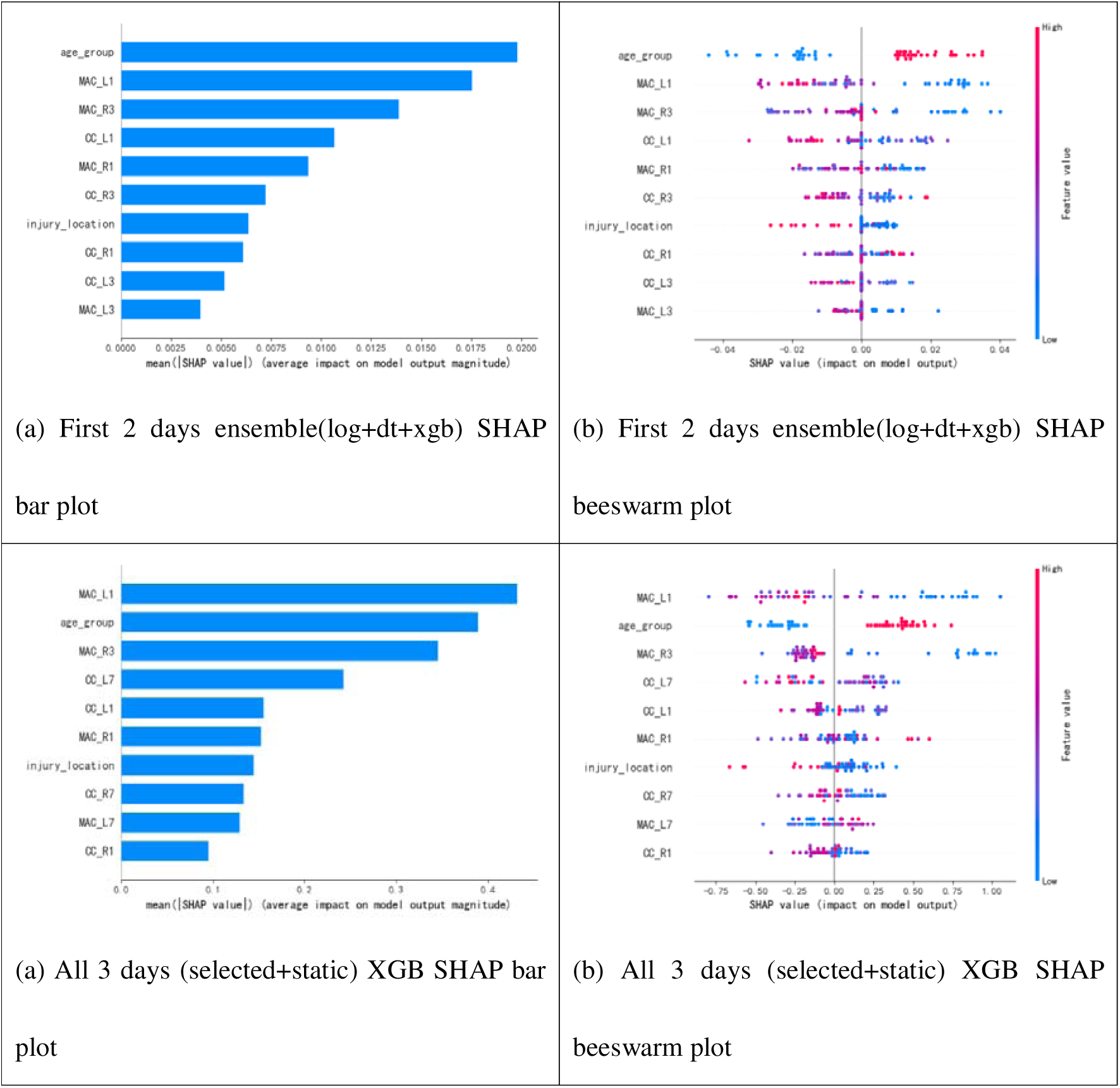
Global SHAP indicator importance and beeswarm plots for the first 2 days ensemble (log+dt+xgb) and 3 days XGBoost models.

For the XGBoost model trained on the dataset including all three time points under the selected indicator setting, MAC_L1 ranks highest in mean absolute SHAP value, followed closely by age group and MAC_R3. Day 7 anthropometric measurements (CC_L7, CC_R7, MAC_L7) also appear among the top contributors alongside early-stage indicators (CC_L1, MAC_R1) and injury location, reflecting the integration of longitudinal information across all three time points. Notably, the beeswarm plot exhibits substantially wider horizontal dispersion than the two-time-point configuration, with SHAP values ranging from approximately −0.75 to +1.0. This broader spread reflects stronger nonlinear interactions and greater between-sample variability in indicator attribution, consistent with XGBoost’s capacity to capture complex indicator dependencies. While the directional interpretation remains coherent—lower anthropometric measurements are generally associated with higher predicted malnutrition risk, the increased dispersion suggests that individual patient trajectories are more heterogeneous when longitudinal information from all three time points is integrated.

Across both configurations, a consistent hierarchy emerges; anthropometric measurements, particularly mid-upper arm and calf circumferences, together with age group, constitute the primary drivers of model predictions. These findings support the clinical interpretability of the learned decision mechanisms and reinforce the importance of core anthropometric indicators in malnutrition risk stratification.

## 5. Discussion

Several limitations should be acknowledged. The study is based on a relatively small cohort, and model development depended on imputation and data augmentation to address missingness and limited sample size. The rarity of some static categories, especially certain injury types and age strata, may also affect the robustness of subgroup inference. In addition, the labels were derived from GLIM-based clinical assessment, which is clinically grounded but still influenced by the availability and quality of bedside data. Future work should therefore focus on external validation in larger multicenter cohorts, calibration analysis, prospective workflow testing, uncertainty quantification of model parameters and data input, and integration with hospital information systems or digital twin platforms. Even with these limitations, the present results support the feasibility of an AI-enabled, low-burden screening toolbox that can assist early nutritional triage and help prioritize patients for more comprehensive assessment and intervention.

The dominance of MAC, CC, and age group in the SHAP analyses is clinically plausible. These anthropometric measures serve as accessible surrogates for peripheral muscle reserve and overall nutritional status, while age reflects vulnerability to catabolic stress, sarcopenia, and reduced physiologic reserve. The consistent presence of early MAC and CC variables among the top predictors in both the ensemble and XGBoost models suggests that the models are not relying on spurious associations but rather learning patterns that align with established clinical understanding of malnutrition in older trauma populations. In this sense, the proposed framework does not replace GLIM; rather, it operationalizes GLIM-oriented risk recognition in a form that is faster, more reproducible, and more amenable to automation.

The results also clarify the value of longitudinal monitoring. Day 1 measurements alone provided only moderate discrimination, whereas adding Day 3 information markedly improved classification performance and stability. By contrast, extending the input window to Day 7 yielded only limited additional benefit, despite stronger discrimination in some configurations, and those gains should be interpreted cautiously because Day 7 had the highest missing-data burden and depended most heavily on imputation. From a clinical perspective, this suggests that the earliest useful assessment window may be the combination of admission and short-interval follow-up measurements, which balances predictive value with practical availability and supports earlier nutritional intervention.

This study demonstrates that clinically practical malnutrition risk assessment in elderly ICU trauma patients does not require an extensive panel of measurements. Across temporal configurations and model classes, bilateral mid-upper arm circumference and calf circumference, combined with a small set of static baseline variables, captured most of the predictive signal and often matched or exceeded the performance of models built with the full set of dynamic indicators. This finding is important for bedside implementation because MAC and CC are simple, inexpensive, and readily obtainable in emergency and critical-care settings, where more comprehensive nutritional assessments are often delayed, incomplete, or operationally difficult.

In practical settings, the models can be implemented within a rolling-window framework, where two or three time-point measurements are used to predict malnutrition status beyond the latest available observation. This design supports its use in elderly ICU patients by collecting longitudinal measurements, thereby providing clinicians and patients with a more informed understanding of nutritional status and its temporal trajectory, with potential implications for integrated treatment planning. While we recommend the ensemble model for analyses based on the first two days of input and XGBoost for those based on the first three days, models trained using other input configurations are also available to end users.

## 6. Conclusion

This study develops a predictive toolbox for malnutrition risk identification in elderly trauma patients using six machine learning models across multiple temporal and indicator configurations. Models incorporating longitudinal measurements generally outperformed those relying on baseline data alone, though Day 7 results should be interpreted with caution, given substantial data missingness and extensive imputation. SHAP analyses consistently identified age_group, MAC_L1, MAC_R3, CC_L1, and MAC_R1 as core predictors across model configurations, reflecting the foundational role of early anthropometric status and age in predicting malnutrition risk.

A selected dynamic indicator set—comprising bilateral MAC and CC measurements combined with static baseline variables—captured most of the predictive information and frequently matched or exceeded the full dynamic indicator set, suggesting that a compact anthropometric representation is sufficient for effective risk stratification.

Tree-based approaches consistently outperformed linear and distance-based classifiers across most configurations. Among individual models, XGBoost demonstrated the most balanced performance, while two-layer ensemble models provided moderate additional improvements by aggregating complementary base learners.

Taking both predictive performance and clinical feasibility into account, the Log+DT+XGB ensemble trained on Day 1 and Day 3 data is recommended as the preferred solution, achieving the highest MCC (0.7044) and accuracy (0.8519) while relying on early-stage measurements that are more readily obtainable in clinical practice. For scenarios requiring full longitudinal integration, the three-time-point XGBoost model remains a robust standalone alternative.

Overall, parsimonious anthropometric indicators, especially early measurements of MAC and CC, combined with age group and tree-based learning approaches, offer an effective and clinically interpretable framework for early malnutrition risk stratification in hospitalized elderly trauma patients. In practice, models trained on two or three consecutive longitudinal measurements can be deployed in a rolling-window framework to predict outcomes at the latest time point within the observation period. Models using single-day input can likewise be retained as reference tools. Collectively, these models are provided as flexible options within the toolbox.

## Declaration of generative AI and AI-assisted technologies in the manuscript preparation process

During the preparation of this work, the author(s) used CHATGPT to conduct English editing in some paragraphs. After using this tool, the author(s) reviewed and edited the content as needed and take(s) full responsibility for the content of the published article.

## Acknowledgements

We appreciate Dr. Hua Jiang of Sichuan Provincial People’s Hospital sharing the dataset with us.

## Data availability

The datasets used and/or analyzed during the current study are available from the corresponding author on reasonable request.

## Code availability

The underlying code for this study and the training/validation datasets are not publicly available but may be made available to researchers on reasonable requests from the corresponding

## Author contributions

Xinhang Wei conducted data analysis and indicator engineering and developed machine learning models.

Xiang Cao and Jianguo Hou assisted with data analysis and model development.

Qi Wang designed, managed, secured funding for the project, and drafted and finalized the article.

